# The Adaptive Physical Activity study in Stroke (TAPAS); A feasibility Sequential Multiple Assignment Randomized Trial

**DOI:** 10.1101/2025.06.10.25329382

**Authors:** Aoife Whiston, Emma Carr, Nathan Cardy, Padraic Rocliffe, Siobhan M. O’Reilly, Daniel Carter, Mairead O’Donoghue, Aine Cronin, James G Bradley, Mike Butler, Liam Glynn, Jane C. Walsh, Cathal Walsh, Kelley Kidwell, Margaret O’Connor, Chris Macey, Lorna Paul, Claire Fitzsimons, Julie Bernhardt, Jon Salsberg, Sara Hayes

## Abstract

**Introduction:** Physical inactivity in people post-stroke increases risk of recurrent stroke and mortality. Personalised physical activity (PA) interventions are recommended. Study designs must be adaptive to individual performance. A Sequential, Multiple Assignment, Randomised Trial (SMART) is a factorial design in a sequential setting used to build effective embedded adaptive interventions. To date, SMARTs have not been used to inform secondary stroke prevention. This study aimed to examine the feasibility of an adaptive, mobile health (mHealth) PA intervention post-stroke, using a SMART.

**Methods:** Community-dwelling people with stroke were randomised to a 12-week exercise (EX) or a lifestyle PA (LPA) intervention delivered via online platform. Six weeks post-randomisation, participants were classified as responders or non-responders according to mean step count/day. Non-responders were re-randomised to switch to a different intervention or augment their original intervention. Responders continued with initial interventions. Primary outcomes included recruitment, retention and adherence rates. Secondary outcomes included PA, sedentary behaviour, fatigue, quality of life, psychological distress, activities of daily living, stroke recurrence and adverse effects. General linear models estimated trends regarding first-stage interventions and non-response strategies, and embedded adaptive interventions were investigated using weighted and replicated regressions.

**Results:** Fifty participants were included, with a mean age of 51.02 years, mean time since stroke of 31.44 months, of which 42% were female and 44% had ischaemic stroke. Recruitment rate was 85%. Retention rates averaged 87% at post-intervention and 80% at the 3-month follow-up. Total mean intervention adherence was 82.16% (SD =18.47%), with trends for higher adherence across LPA intervention components. No significant differences were observed for secondary outcomes comparing first-stage interventions. Positive trends were observed for switching interventions and potential benefits of embedded adaptive interventions.

**Conclusion:** This feasibility SMART provides first-in-class empirical data on the design of an adaptive mHealth PA intervention post-stroke. A definitive SMART would be feasible.

**Clinical Trials Registration:** NCT05606770

## Introduction

Stroke remains the second leading cause of death and disability globally (1), despite advances in stroke interventions and rehabilitation. Meta-analytic evidence demonstrates that the 1-year and 5-year risk of recurrent stroke is 11.1% and 26.4%, respectively (2). People with stroke who are physically inactive have more than three times the risk of recurrent stroke as their peers (3). Additionally, more than 50% of people with stroke have impaired walking and decreased independence at 6-12 months post-stroke (4). This is both a consequence of and risk factor for physical inactivity (4) and puts people with stroke at an increased risk of recurrent stroke or death (4, 5, 6). This well-established link between physical inactivity, disability and stroke recurrence underscores the critical need to enhance physical activity (PA) levels among people with stroke.

PA is defined as body movement produced by skeletal muscles resulting in energy expenditure (7). Interventions to improve PA levels in people post-stroke are often multi-component and include treatments to increase lifestyle PA (e.g., take more steps during daily tasks) and treatments to promote structured exercise (e.g., engage in bouts of moderate-to-vigorous structured exercise) (8). A Cochrane review of 75 trials examined the effectiveness of structured exercise in people with stroke and found moderate improvements in disability outcomes (9). However, these benefits were short-term due to little or no emphasis on modifying everyday behaviours surrounding PA levels. Reducing recurrent strokes and improving functional outcomes often requires sustained positive behaviours, supported by theory-driven behaviour change interventions (10). For example, a recent randomised controlled trial involving 250 people with stroke evaluated a high intensity walking program (FAST), a step activity monitoring behavioural intervention (SAM), and their combination (FAST + SAM) and found that PA gains were greater in programmes that included behaviour change strategies (e.g., skills coaching and goal setting) (11). Similarly, a systematic review of 11 studies post-stroke showed interventions to promote PA should incorporate PA-specific tailored counselling based on sound behavioural theory vs. general advice only (12). These tailored behaviour-change interventions can now be delivered via mHealth technologies-mobile-based tools that support medical and public health practices using devices like smartphones, patient monitors, and other wireless technologies (13).

Interest has grown in mHealth as a cost-effective, scalable and accessible tool for delivering personalised interventions post-stroke (14). A recent scoping review of 17 studies found that SMS and smartphone applications, used alone or together, are the most common mHealth tools for supporting lifestyle PA changes in secondary stroke prevention (15). While a Cochrane review of four studies found inconsistent evidence for the effectiveness of wearable activity monitors for enhancing PA after stroke, this was attributed to the limited use of specific behaviour change strategies across these interventions (16). Conversely, a systematic review of PA interventions across a variety of populations emphasised that, due to the inherent intervention complexity, which often require tailoring in terms of type, intensity and dosage, adaptive approaches incorporating mHealth technologies, are essential to effectively address the diverse needs and outcomes experienced by people after stroke (17). Thus, the usability and key features of mHealth to increase PA and promote secondary prevention post-stroke remain unclear.

Prior to the design of the TAPAS trial, a qualitative study was conducted to explore the perspectives of people with stroke, caregivers and healthcare professionals regarding the development and implementation of an adaptive, personalised mHealth intervention aimed at PA after stroke (18). The study identified three main themes: essential features of an mHealth intervention, strategies for effective delivery, and barriers to development and use. Across all stakeholder groups, there was strong consensus that an adaptive mHealth intervention to support PA post-stroke would be highly beneficial, particularly following discharge from acute care.

An embedded adaptive intervention (EAI) is a set of therapeutic strategies that are used in stages and the selection of the intervention at each stage is based on defined decision rules. Typical adaptations include augmenting an ongoing treatment or switching to another treatment. These decisions are made in response to changes in the person’s status, such as a person’s response or non-response to treatment. The person experiences an EAI as a sequence of personalised treatments (19). While this is reflective of rehabilitation practices post-stroke, there is a lack of empirical data on the optimum sequence of these treatments in the existing evidence base. Therefore, study designs used in the evaluation of PA interventions must be adaptive to individual response (17). There has been an increased interest by stroke researchers in the potential for the use of innovative trials designs, in response to acknowledged difficulties relating to recruitment, administrative burden and cost in completing stroke trials (20). A Sequential, Multiple Assignment, Randomised Trial (SMART) is a type of multistage, factorial randomised trial, in which some or all participants are randomised at two or more decision points. Whether a person is randomised at the second or a later decision point, and the available treatment options, may depend on their response to prior treatment-reflective of an EAI (19). SMARTs facilitate the evaluation of EAIs by systematically testing different sequences of intervention options.

The current study is the first to examine the feasibility of an adaptive, mHealth PA intervention for community-dwelling people with stroke, using a SMART design. While pre-specified feasibility targets were not set, to aid hypothesis testing, findings are reported in the context of the existing evidence base regarding recruitment, retention and adherence rates among previous trials of PA interventions post-stroke. The trends in clinical efficacy of the EAIs were also examined. In accordance with established guidelines for the design, implementation, and analysis of pilot SMARTs (21), this study was not designed to conduct statistical tests for differences in clinical outcomes across intervention conditions.

## Methods

The data that support the findings of this study are available from the corresponding author upon reasonable request. The TAPAS trial was a multisite assessor-blinded feasibility SMART. In the first stage, participants were randomised to structured exercise (EX) or lifestyle PA (LPA) interventions (Table 1 and 2). Common to both intervention components was the provision of a Fitbit Inspire 2 and daily step count goals. Participants completed a 12-week intervention. At Week 6, the research team remotely determined if participants were deemed responders or non-responders to their initial intervention assignment based on a *tailoring variable*: an objective measurement of their daily PA step count and a non-responder *decision rule*. Per this rule, participants were classified as non-responders if a) their average 7-day step count did not meet the short-term goal of 5% more than the 7-day average from their previous week, for two of the three weeks in Weeks 4 to 6; or b) they failed to wear their Fitbit or did not have a valid day (i.e., wore the Fitbit a minimum of three of seven days in Weeks 4 to 6, with either 10 hours of wear time for that day or exceed the target step count for that day). The choice of 5% weekly increments in step count targets was based on feasible step count goals used in previous trials among ambulatory people with stroke (22,23). In the second stage, non-responders to initial treatments were randomly assigned to one of the four alternative treatments, dependent on their initial treatment: a) continue EX augmented with LPA, b) switch to EX alone, c) LPA augmented with EX, d) switch to LPA alone. Responders continued with their original treatment for Weeks 6 to 12. The EAIs are outlined in **Table 3**, and an overview of the participant flow throughout the SMART is presented in **Figure 1**.

**Figure 1.**
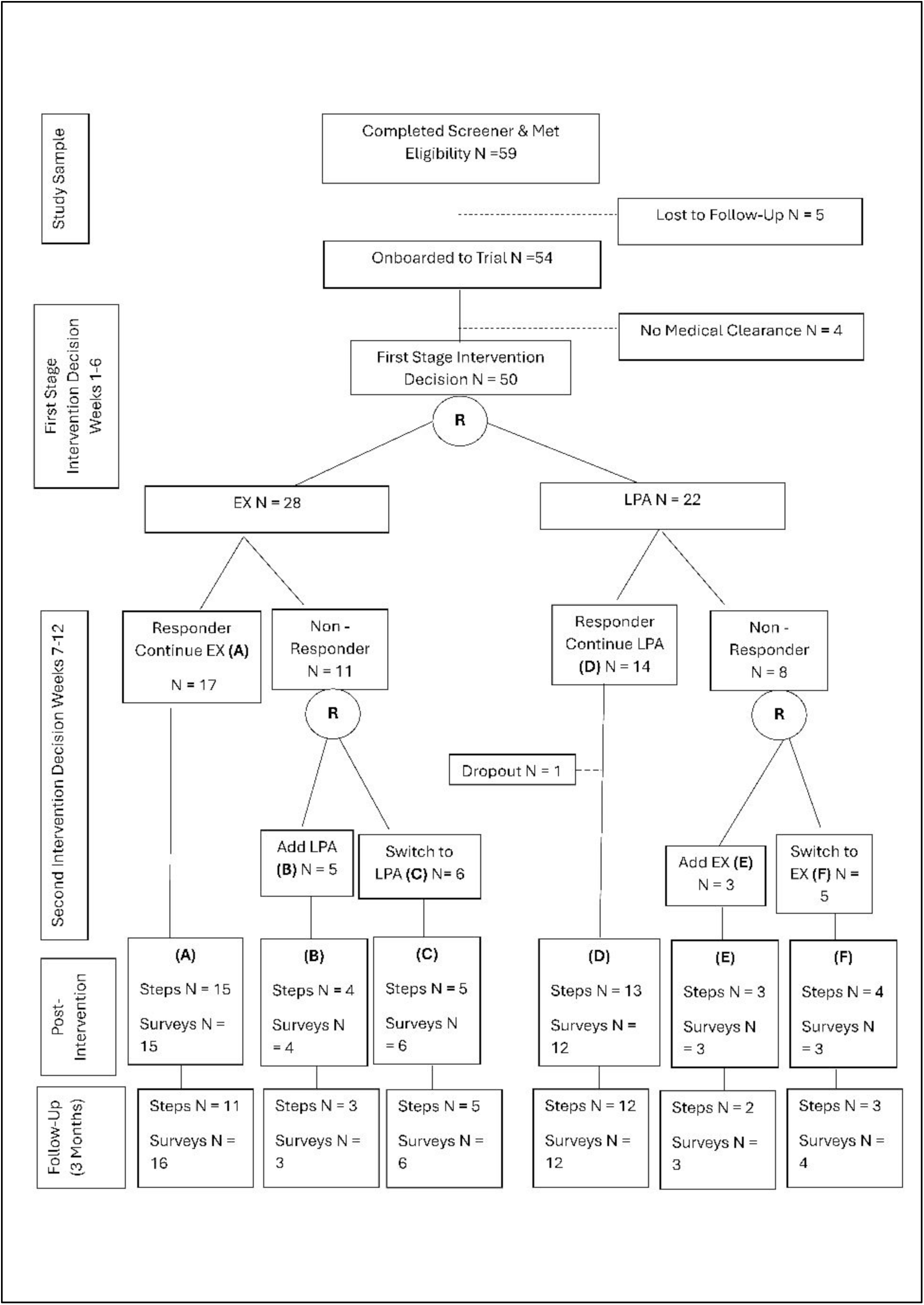
Participant Flow Diagram.

**Table 1.**
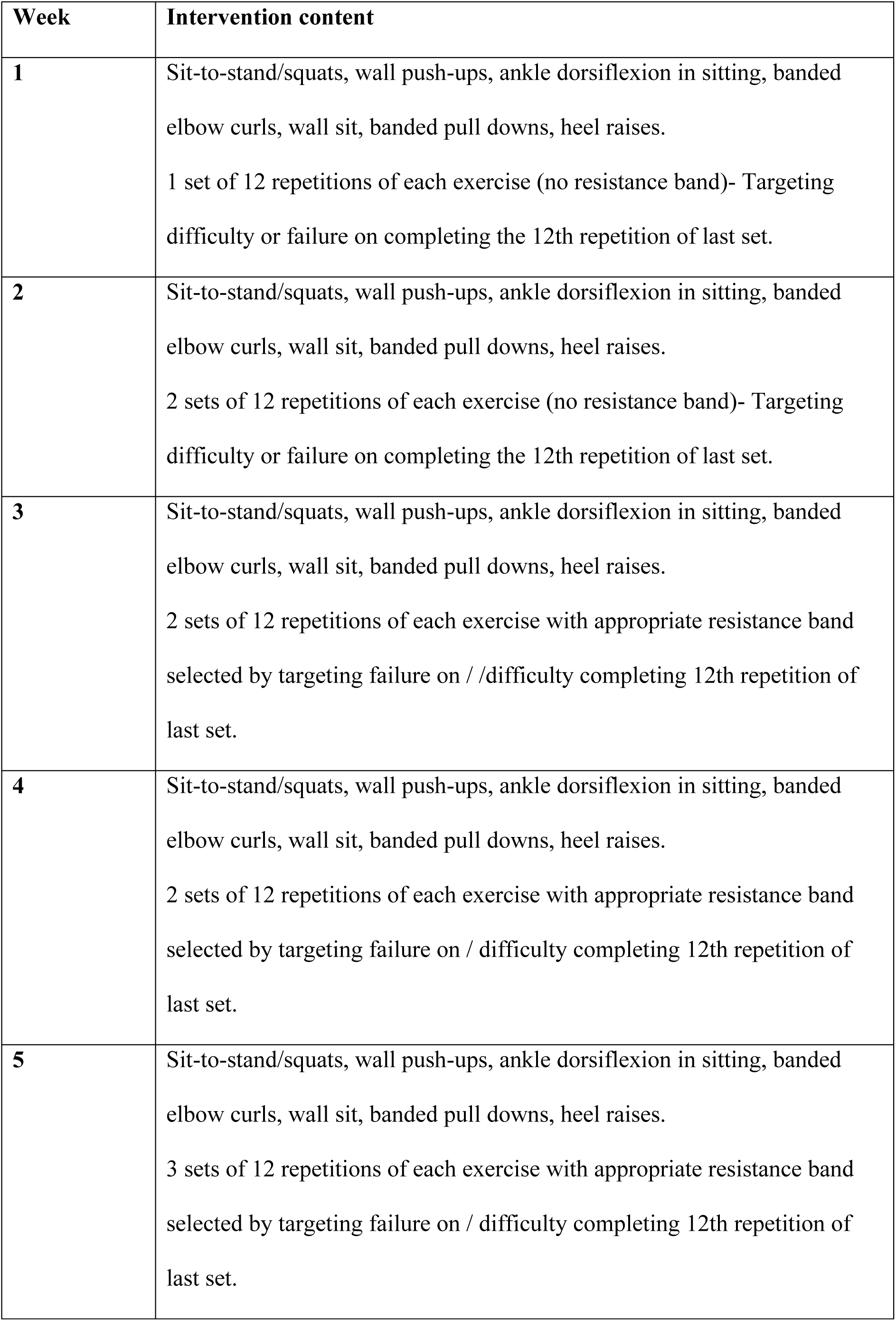
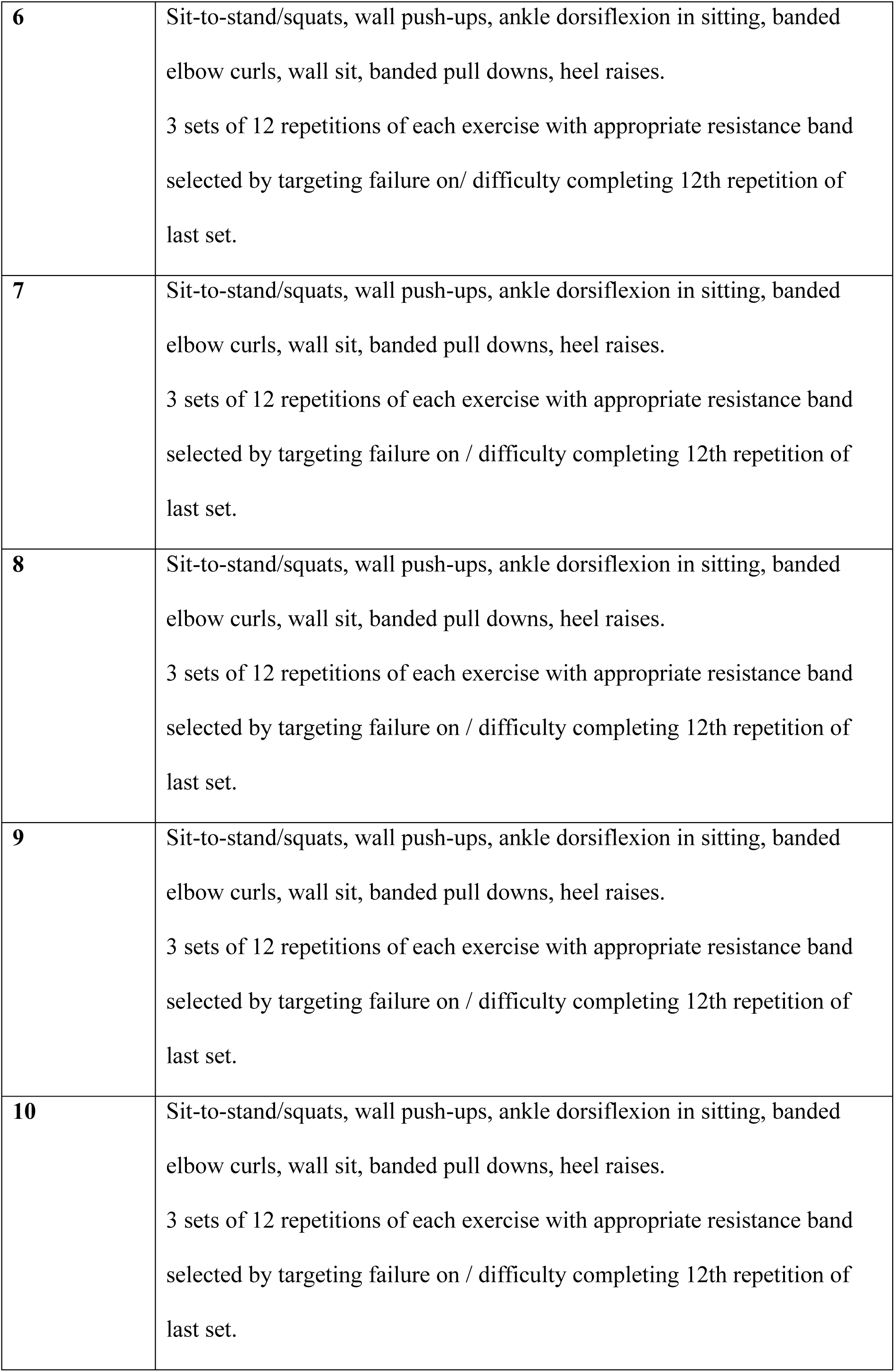
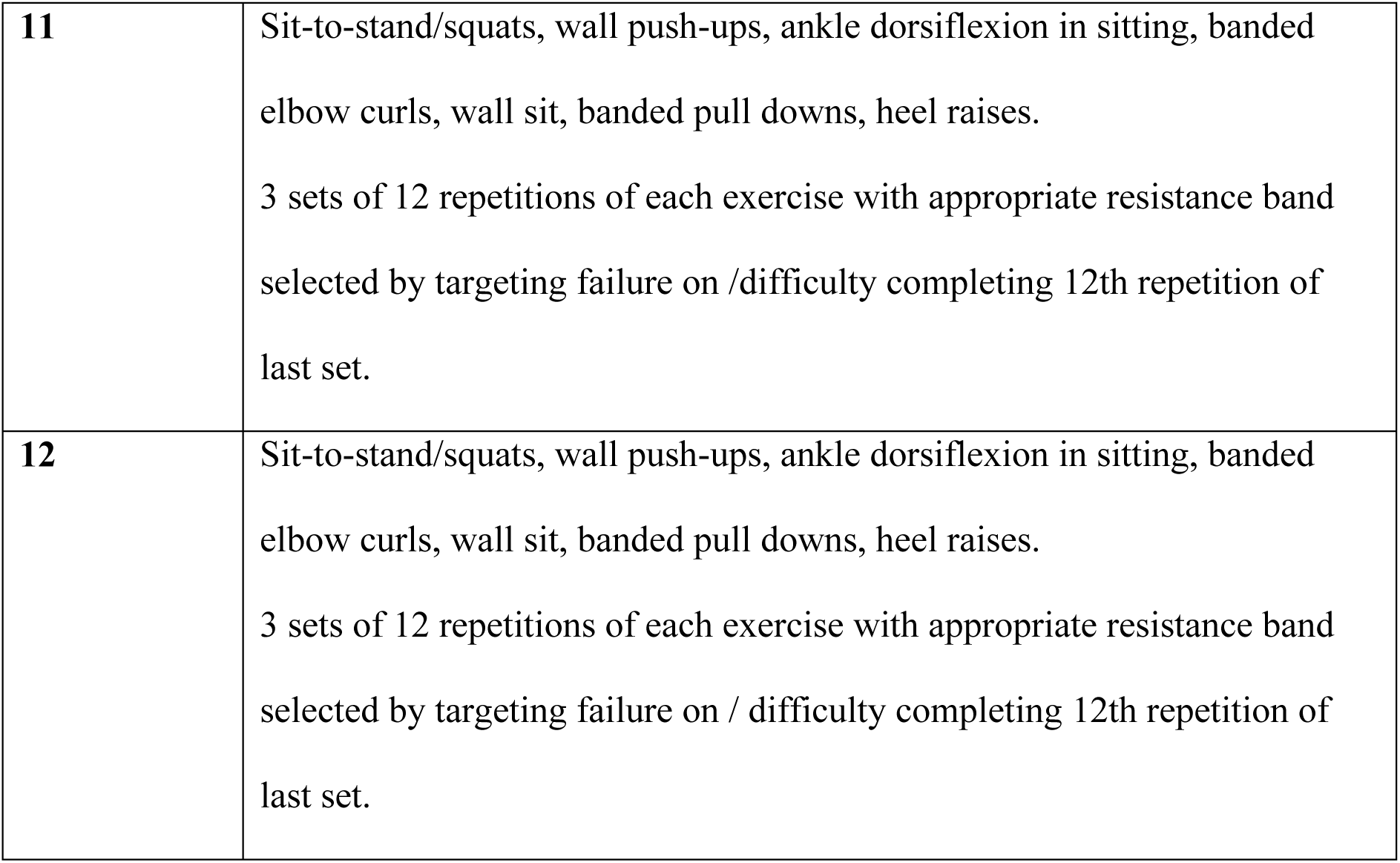
Overview of Structured Exercise Program (EX).

| **Week** | **Intervention content** |
| --- | --- |
| **1** | Sit-to-stand/squats, wall push-ups, ankle dorsiflexion in sitting, banded elbow curls, wall sit, banded pull downs, heel raises.<br>1 set of 12 repetitions of each exercise (no resistance band)- Targeting difficulty or failure on completing the 12th repetition of last set. |
| **2** | Sit-to-stand/squats, wall push-ups, ankle dorsiflexion in sitting, banded elbow curls, wall sit, banded pull downs, heel raises.<br>2 sets of 12 repetitions of each exercise (no resistance band)- Targeting difficulty or failure on completing the 12th repetition of last set. |
| **3** | Sit-to-stand/squats, wall push-ups, ankle dorsiflexion in sitting, banded elbow curls, wall sit, banded pull downs, heel raises.<br>2 sets of 12 repetitions of each exercise with appropriate resistance band selected by targeting failure on / /difficulty completing 12th repetition of last set. |
| **4** | Sit-to-stand/squats, wall push-ups, ankle dorsiflexion in sitting, banded elbow curls, wall sit, banded pull downs, heel raises.<br>2 sets of 12 repetitions of each exercise with appropriate resistance band selected by targeting failure on / difficulty completing 12th repetition of last set. |
| **5** | Sit-to-stand/squats, wall push-ups, ankle dorsiflexion in sitting, banded elbow curls, wall sit, banded pull downs, heel raises.<br>3 sets of 12 repetitions of each exercise with appropriate resistance band selected by targeting failure on / difficulty completing 12th repetition of last set. |
| **6** | Sit-to-stand/squats, wall push-ups, ankle dorsiflexion in sitting, banded elbow curls, wall sit, banded pull downs, heel raises.<br>3 sets of 12 repetitions of each exercise with appropriate resistance band selected by targeting failure on/ difficulty completing 12th repetition of last set. |
| **7** | Sit-to-stand/squats, wall push-ups, ankle dorsiflexion in sitting, banded elbow curls, wall sit, banded pull downs, heel raises.<br>3 sets of 12 repetitions of each exercise with appropriate resistance band selected by targeting failure on / difficulty completing 12th repetition of last set. |
| **8** | Sit-to-stand/squats, wall push-ups, ankle dorsiflexion in sitting, banded elbow curls, wall sit, banded pull downs, heel raises.<br>3 sets of 12 repetitions of each exercise with appropriate resistance band selected by targeting failure on / difficulty completing 12th repetition of last set. |
| **9** | Sit-to-stand/squats, wall push-ups, ankle dorsiflexion in sitting, banded elbow curls, wall sit, banded pull downs, heel raises.<br>3 sets of 12 repetitions of each exercise with appropriate resistance band selected by targeting failure on / difficulty completing 12th repetition of last set. |
| **10** | Sit-to-stand/squats, wall push-ups, ankle dorsiflexion in sitting, banded elbow curls, wall sit, banded pull downs, heel raises.<br>3 sets of 12 repetitions of each exercise with appropriate resistance band selected by targeting failure on / difficulty completing 12th repetition of last set. |
| **11** | Sit-to-stand/squats, wall push-ups, ankle dorsiflexion in sitting, banded elbow curls, wall sit, banded pull downs, heel raises.<br>3 sets of 12 repetitions of each exercise with appropriate resistance band selected by targeting failure on /difficulty completing 12th repetition of last set. |
| **12** | Sit-to-stand/squats, wall push-ups, ankle dorsiflexion in sitting, banded elbow curls, wall sit, banded pull downs, heel raises.<br>3 sets of 12 repetitions of each exercise with appropriate resistance band selected by targeting failure on / difficulty completing 12th repetition of last set. |

**Table 2.** Overview of Lifestyle PA Component (LPA).

| **Week** | **Intervention content** |
| --- | --- |
| **1** | Video: LPA introduction<br>Video: What is PA?<br>Video: How do I increase my PA?<br>Weekly text: Individualised daily step count target<br>Step count strategies offered via multiple choice<br>Scripted phone call/texts<br>Problem solving |
| **2** | Video: Benefits of PA<br>Weekly text: Individualised daily step count target<br>Step count strategies offered via multiple choice<br>Scripted phone call/texts<br>Problem solving |
| **3** | Video: Is PA safe after stroke?<br>Weekly text: Individualised daily step count target<br>Step count strategies offered via multiple choice<br>Scripted phone call/texts:<br>Problem solving |
| **4** | Video: Hear from a physiotherapist<br>Weekly text: Individualised daily step count target<br>Step count strategies offered via multiple choice<br>Scripted phone call/texts<br>Problem solving |
| **5** | Video: Social connection and PA<br>Weekly text: Individualised daily step count target<br>Step count strategies offered via multiple choice<br>Scripted phone call/texts<br>Problem solving |
| **6** | Video: Barriers and facilitators to PA<br>Video: Harmful effects of no PA<br>Weekly text: Individualised daily step count target<br>Step count strategies offered via multiple choice<br>Scripted phone call/texts<br>Problem solving |
| **7-12** | Repeat Week 1-6 |

**Table 3.** SMART Design: Stages of Intervention and EAIs.

|  |  |  |  |
| --- | --- | --- | --- |
| **Embedded** | **First Stage** | **Tailoring** | **Second Stage** |

| Adaptive Intervention (EAI) | Intervention | Variable - Responder | Intervention |
| --- | --- | --- | --- |
| 1 | EX | Yes | EX |
|  |  | No | EX+LPA |
| 2 | EX | Yes | EX |
|  |  | No | LPA |
| 3 | LPA | Yes | LPA |
|  |  | No | LPA+EX |
| 4 | LPA | Yes | LPA |
|  |  | No | EX |

EX = Structured Exercise Intervention; LPA = Lifestyle Physical Activity Intervention.

The a-priori determined primary outcome and endpoint was change in PA, measured by mean steps/day assessed for one week at baseline, post-intervention, and at 3-month follow-up (24). However, due to time constraints, a feasibility SMART was conducted. This change in protocol was enacted in August 2023, before recruitment began for the trial. It was based on the research teams experience of a relatively low recruitment rate in a different trial in the same recruitment centres, and availability of resources for the TAPAS trial. Feasibility was measured using recruitment, retention and adherence rates and adverse events and the current trial is reported as a feasibility SMART. The original study protocol has been published (24). Here, we report on the updated feasibility protocol (Clinical Trials Registration: NCT05606770) and is reported in according to CONSORT extension to pilot and feasibility trials (25). A formal sample size calculation was not conducted. A target sample size of approximately 40 participants was pragmatically selected based on common practice in feasibility trials (26, 27).

### Participants

Participants were recruited from the community through the following sites in Ireland: University Hospital Limerick; St. Camillus’ Community Hospital, Limerick; St. Ita’s Hospital, Limerick; and the Irish Heart Foundation. Participants were enrolled from October 2023 to March 2024. Participants provided written informed consent and met the inclusion criteria as follows: (1) aged 18 years or older; (2) living in the community; (3) had a clinician-confirmed stroke; (4) were no longer receiving inpatient, outpatient or community-based PA rehabilitation; (5) were independently mobile (with or without a mobility aid); (6) able to express their basic needs verbally; (7) had access to relevant technology to enable mHealth intervention delivery; (8) had cognitive capacity to provide informed consent; and (9) had medical clearance for participation in the TAPAS programme. Exclusion criteria included: (1) contraindications for undertaking PA (e.g., safety, presence of unstable heart disease).

### Randomisation and Masking

Participants were randomly assigned to one of two first-stage intervention components: EX or LPA, following screening and baseline testing. A simple equal allocation randomisation at the individual level using a computer-generated randomised list, which was concealed from researchers involved in trial enrolment, was used. At Week 6, participants were classified as either responders or non-responders according to their step count change or fitbit wear. Participants classified as responders continued in their respective groups until the end of the 12-week intervention. Participants classified as non-responders were re-randomised to alternative treatment options for the second six-week period (Figure 1). Both randomisations were performed by a statistician on the research team. Outcome assessors and data analysts were blinded to all treatment allocations.

### Procedure

Participants were enrolled to the trial by AW in an in-person meeting at the University of Limerick. Participants were given a Fitbit Inspire 2 and a tutorial and written instructions on its use and were also shown how to log into and navigate the specifically-designed online platform (TAPAS application).

### Interventions

There were two components which targeted increased PA in the current intervention: (EX and LPA). The content of both interventions has been outlined in detail in the protocol paper (24) and is further outlined in Tables 2 and 3. Details of the behaviour change techniques identified within the Fitbit Inspire 2 are described previously (24). The Fitbit Inspire 2 was used to measure daily step count among participants and inform the step count goal in both intervention components. Individualized baseline and continuous step count data for participants were used to assign daily step count goals each week. All participants who achieved their daily step count goal from the previous week were assigned a new daily step count goal each week calculated by adding 5% to the 7-day average from their previous week. Otherwise, their daily step count goal remained unchanged from week to week.

Participants assigned to EX were provided with twice weekly circuit strengthening exercise sessions, delivered through the online TAPAS application. Each class included a warm-up and cool down. The LPA component also delivered on the TAPAS application, was developed using the Behaviour Change Wheel (BCW) Guide to Designing Interventions and is underpinned by the COM-B model of behavior change (28). The aim of the LPA component was to increase the capability, opportunity, and motivation of participants to reach their daily step count goals.

### Outcomes

All participants completed online demographic and clinical self-report questionnaires, in addition to objective step count measurement at baseline, post-intervention, and at 3-month follow-up time-points.

#### Primary outcomes

Feasibility was measured using recruitment, retention, and adherence rates and adverse events. Pre-specified feasibility targets were not identified, however, the findings in relation to feasibility outcomes are discussed in the context of the existing evidence base. Recruitment rate was calculated as a percentage, with the number of people enrolled divided by the number of eligible participants. Retention was calculated as the percentage of participants who, after enrolment, completed the pre-intervention outcome measures and post-intervention outcome measures. We reported this according to objective step count data and self-report questionnaires. Adherence to the intervention was measured by calculating the percentage of treatment sessions completed by participants. The denominator for this calculation differed according to the EAI. Processing time for enrolling participants was measured in days. Adverse events were assessed using self-report-questions.

#### Secondary outcomes

The psychometric properties of the included secondary outcome measures have been previously outlined (24). Mean steps/day over 7-days were measured using the Fitbit Inspire 2 on the non-paretic limb. A minimum of four days of steps data was required to calculate a weekly average, aligning with previous research (29, 30). Sedentary behaviour was measured using the Sedentary Behaviour Questionnaire (SBQ) (31). Fatigue was measured using the 7-item self-reported Fatigue Severity Scale (32). Quality of Life was measured using the Stroke Specific Quality of Life scale (SS-QOL) (33) and the EQ-5D-5L-index (34). Psychological distress was measured using the Hospital Anxiety and Depression Scale (HADS) (35). Activities of daily living were measured using the 11-item Re-Integration into Normal Living Index (RNLI) (36). Cognitive function was measured using the Cognitive Assessment Scale for Stroke Patients (CASP) (37) at baseline only.

### Statistical Analysis

To address feasibility aims, we reported descriptive statistics relating to proportions and confidence intervals. Baseline characteristics according to initial group allocation were also presented. To estimate average trends of clinical efficacy related to comparing first-stage interventions (EX vs. LPA), general linear models were estimated at post-intervention and 3-month follow-up, controlling for baseline scores on each secondary outcome. Second, to identify the most effective second-stage strategy for non-responders-switching versus augmenting interventions, the data were subset to consider only non-responders and general linear models were estimated at post-intervention and 3-month follow-up, controlling for baseline scores on each secondary outcome and first-stage intervention effects. Finally, to estimate trends of the four EAIs, a weighted and replicated regression approach was applied to all secondary outcomes. Given the simple equal randomisation structure, participants were randomised to first-stage interventions (EX vs. LPA), with responders continuing their intervention, and non-responders further randomised to stage-two interventions (switch versus augment treatment). To adjust for design imbalances, known probability of randomisation weights were assigned-responders received a weight of 2 (1/0.5) and non-responders a weight of 4 (1/0.5*0.5). As responders are consistent with two EAIs, their data were replicated. This enabled their data to inform the estimation of both EAIs. To account for the replicated data and variability in weights, a generalized estimating equation (GEE) model with robust standard errors was fit to the data (38). The same analytic approach was applied to examine trends in total intervention adherence-adherence to Fitbit and TAPAS application, for the 12-week intervention across first-stage interventions, non-responder strategies and EAIs. No interim analyses or stopping guidelines were planned or conducted. No post hoc analyses were conducted. All missing data were handled using listwise deletion.

All analyses were prespecified in the study protocol unless otherwise stated (24, Clinical Trials Registration: NCT05606770).

### Patient and Public Involvement

A public and patient involvement (PPI) panel was established including people with stroke and carers, before the initiation of this trial. The panel consisted of sixteen people-eight people with stroke, one carer, three healthcare professionals, and three stroke researchers. The panel met four to five times annually for the duration of this trial and were involved in a meaningful way in the design, delivery, evaluation, and dissemination of the trial (39).

### Ethics

Ethical approval was granted by the Faculty of Education and Health Sciences Research Ethics Committee at the University of Limerick [2022 02 15 EHS (OA)] and the HSE Mid-Western Ethics Committee [REC Ref: 026/2022].

## Results

### Baseline Characteristics

Table 4 demonstrates baseline participant characteristics. Participants included 21 (42%) females and had a mean age of 51.02, standard deviation (SD) = 14.99. The majority (74%, n=37) of the participants were married or cohabiting. Thirty-four percent (n =17) were unable to work due to health reasons, 38% (n=19) were in full-time or part-time employment, 20% (n=10) were retired, and 6% (n=3) were unemployed. The mean time since stroke was 31.43 (SD=42.22) months and 44% (n=22) of the sample reported ischaemic stroke subtype.

**Table 4.**
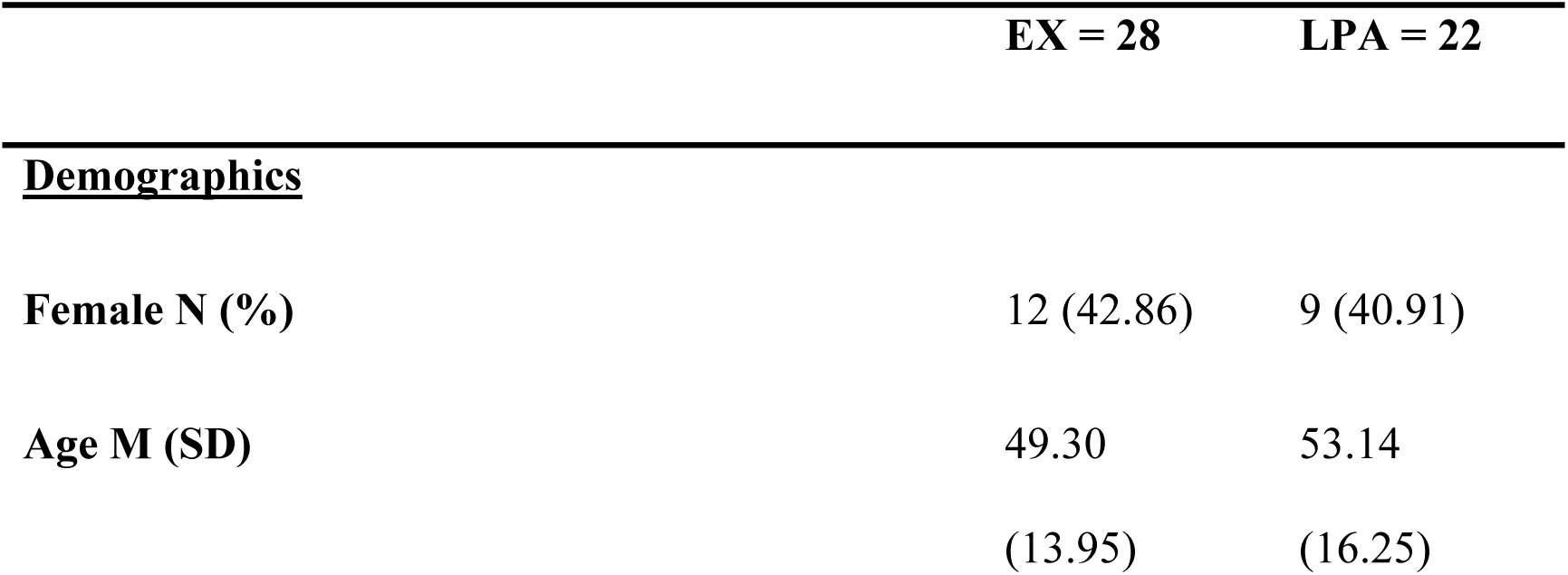
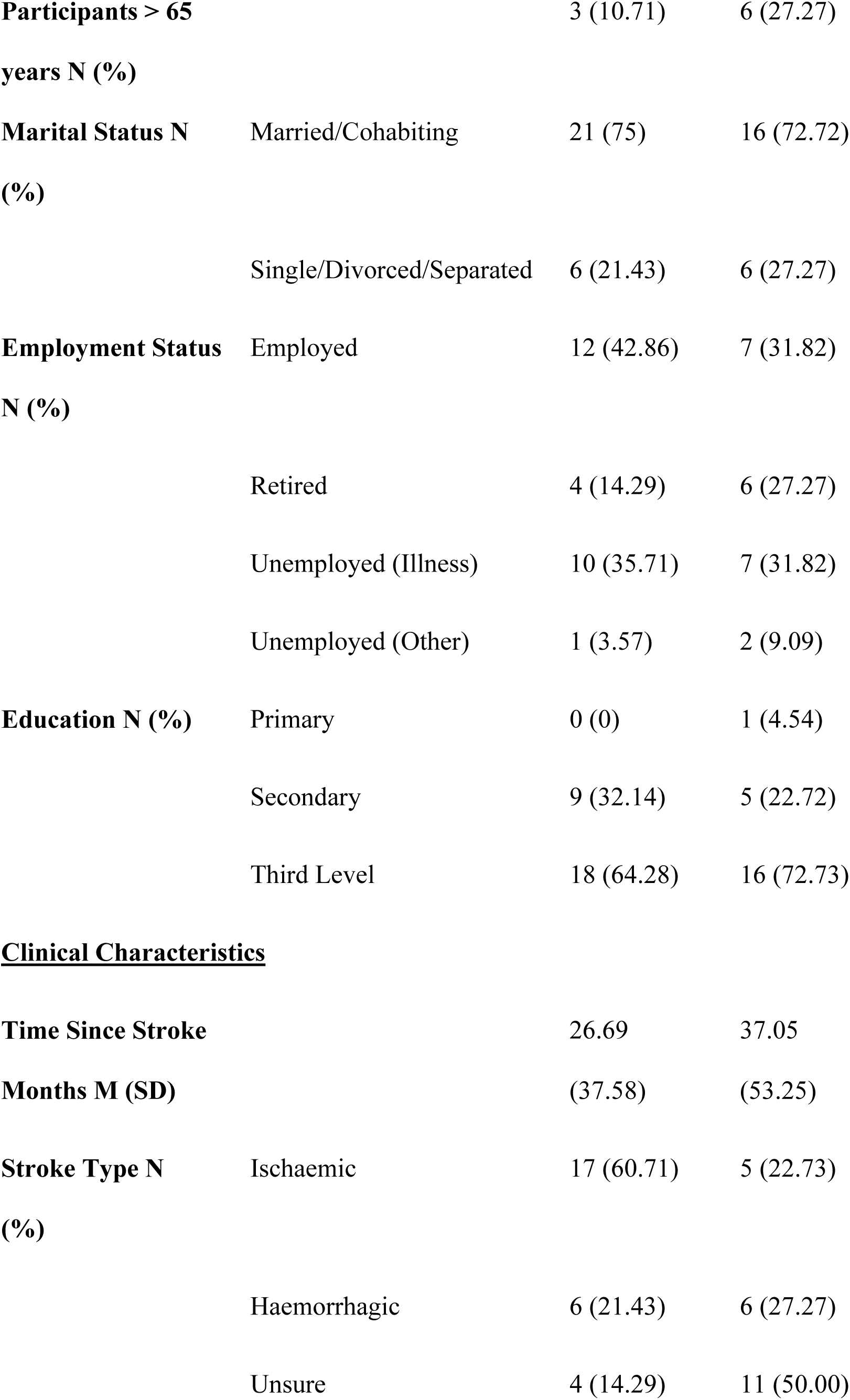
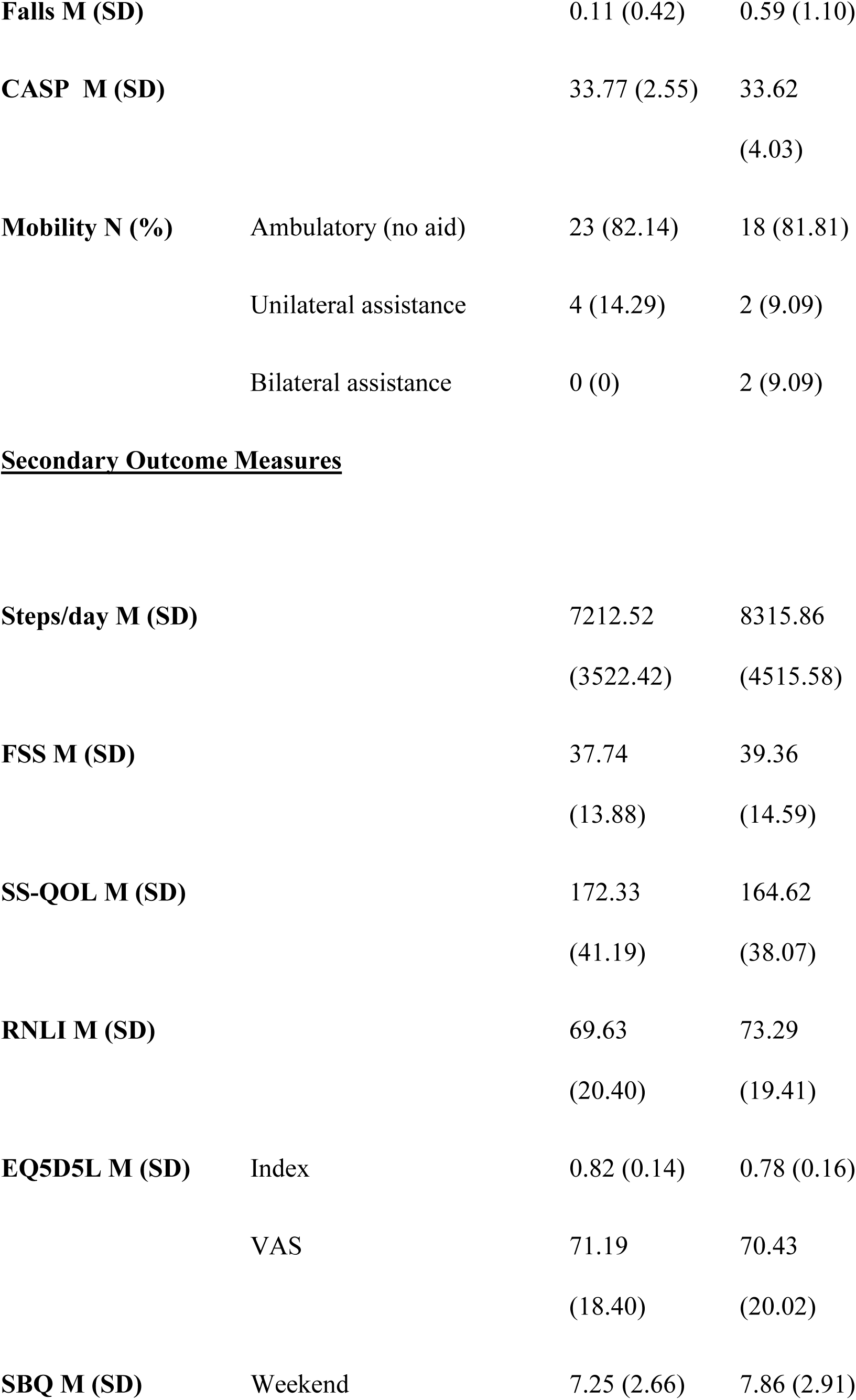
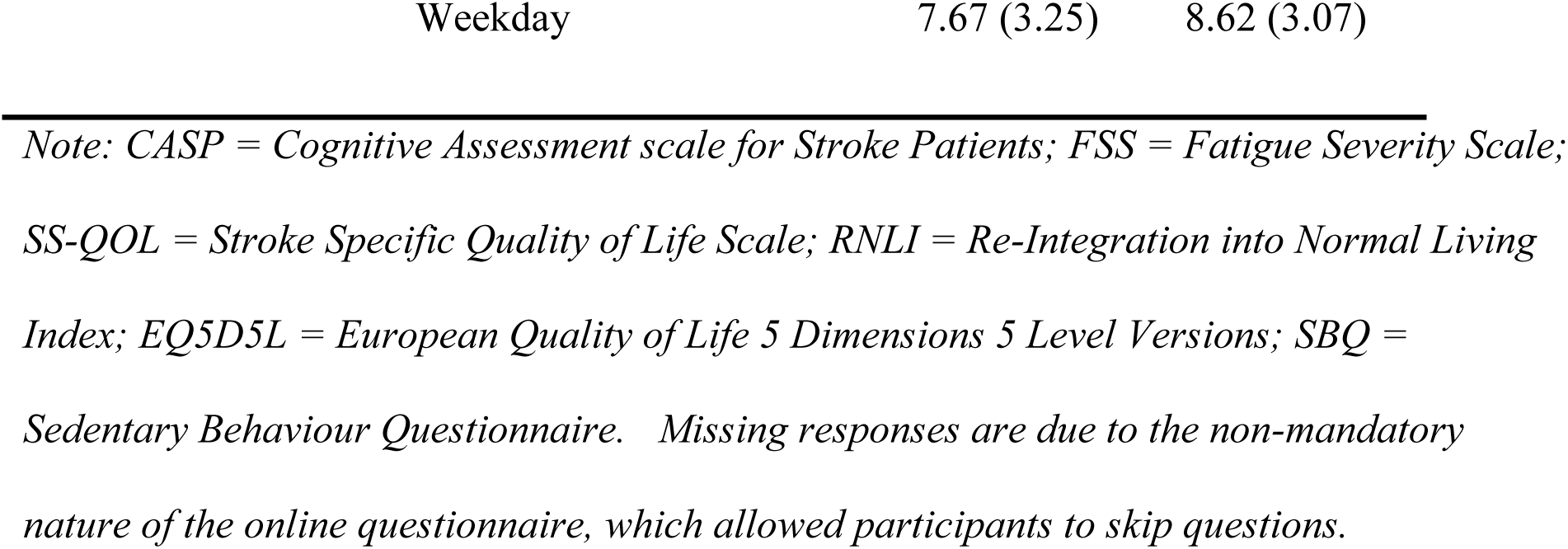
Participant Demographics at Baseline (n=50).

|  | EX = 28 | LPA = 22 |
| --- | --- | --- |
| **<u>Demographics</u>** |  |  |
| **Female N (%)** | 12 (42.86) | 9 (40.91) |
| **Age M (SD)** | 49.30<br>(13.95) | 53.14<br>(16.25) |

|  |  |  |  |
| --- | --- | --- | --- |
| **Participants > 65**<br>**years N (%)** |  | 3 (10.71) | 6 (27.27) |
| **Marital Status N**<br>**(%)** | Married/Cohabiting | 21 (75) | 16 (72.72) |
|  | Single/Divorced/Separated | 6 (21.43) | 6 (27.27) |
| **Employment Status**<br>**N (%)** | Employed | 12 (42.86) | 7 (31.82) |
|  | Retired | 4 (14.29) | 6 (27.27) |
|  | Unemployed (Illness) | 10 (35.71) | 7 (31.82) |
|  | Unemployed (Other) | 1 (3.57) | 2 (9.09) |
| **Education N (%)** | Primary | 0 (0) | 1 (4.54) |
|  | Secondary | 9 (32.14) | 5 (22.72) |
|  | Third Level | 18 (64.28) | 16 (72.73) |
| **<u>Clinical Characteristics</u>** |  |  |  |
| **Time Since Stroke**<br>**Months M (SD)** |  | 26.69<br>(37.58) | 37.05<br>(53.25) |
| **Stroke Type N**<br>**(%)** | Ischaemic | 17 (60.71) | 5 (22.73) |
|  | Haemorrhagic | 6 (21.43) | 6 (27.27) |
|  | Unsure | 4 (14.29) | 11 (50.00) |
| **Falls M (SD)** |  | 0.11 (0.42) | 0.59 (1.10) |
| **CASP M (SD)** |  | 33.77 (2.55) | 33.62<br>(4.03) |
| **Mobility N (%)** | Ambulatory (no aid) | 23 (82.14) | 18 (81.81) |
|  | Unilateral assistance | 4 (14.29) | 2 (9.09) |
|  | Bilateral assistance | 0 (0) | 2 (9.09) |

**Secondary Outcome Measures**
|  |  |  |  |
| --- | --- | --- | --- |
| **Steps/day M (SD)** |  | 7212.52<br>(3522.42) | 8315.86<br>(4515.58) |
| **FSS M (SD)** |  | 37.74<br>(13.88) | 39.36<br>(14.59) |
| **SS-QOL M (SD)** |  | 172.33<br>(41.19) | 164.62<br>(38.07) |
| **RNLI M (SD)** |  | 69.63<br>(20.40) | 73.29<br>(19.41) |
| **EQ5D5L M (SD)** | Index | 0.82 (0.14) | 0.78 (0.16) |
|  | VAS | 71.19<br>(18.40) | 70.43<br>(20.02) |
| **SBQ M (SD)** | Weekend | 7.25 (2.66) | 7.86 (2.91) |

*Note: CASP = Cognitive Assessment scale for Stroke Patients; FSS = Fatigue Severity Scale;* *SS-QOL = Stroke Specific Quality of Life Scale; RNLI = Re-Integration into Normal Living* *Index; EQ5D5L = European Quality of Life 5 Dimensions 5 Level Versions; SBQ =* *Sedentary Behaviour Questionnaire. Missing responses are due to the non-mandatory* *nature of the online questionnaire, which allowed participants to skip questions.*

Participants had a mean score of 33.70 (SD=3.23) on the CASP, indicating good cognitive function. For mobility, 82% (n=41) were ambulatory without aid, 12% (n=6) were ambulatory with unilateral assistance, and 2% (n=1) were ambulatory with bilateral assistance. The mean number of falls reported in the last three months was 0.33 (SD=0.83).

### Primary outcomes-feasibility

As shown in Figure 1, of those who expressed interest in the trial, 93.65% (n=59) were eligible for inclusion, 91.53% (n=54) were enrolled to the trial and 84.75% (n=50) started the intervention. There was one formal drop-out due to work commitments. Retention rates at post-intervention were 88% for objective PA measurement and 86% for self-report questionnaires. At the three-month follow-up, retention rates were 72% for mean steps/day and 88% for questionnaires. Processing time from initial contact to trial enrolment averaged 21.08 (SD =28.94) days. Adverse events recorded at post-intervention included three self-reported falls. At the three-month follow-up, four recurrent strokes and three falls were also reported. Total intervention adherence was an average of 82.16% (SD =18.47%). Generally, adherence to use of the Fitbit showed higher adherence (M=90.69, SD =20.29) compared to adherence to use of the online platform-TAPAS application adherence (M= 56.29, SD =31.14). No significant differences were observed across adherence rates regarding first-stage treatment assignment (EX vs. LPA), non-responder strategies (switching versus augmenting treatment) and EAIs. However, marginal means indicated positive trends favouring LPA (vs. EX) for a first-stage treatment assignment and EAI3 – first-stage treatment of LPA with a non-response strategy to augment with EX, regarding intervention adherence. See Table 5 for an overview of estimated marginal means of adherence across SMART outcomes.

**Table 5.**
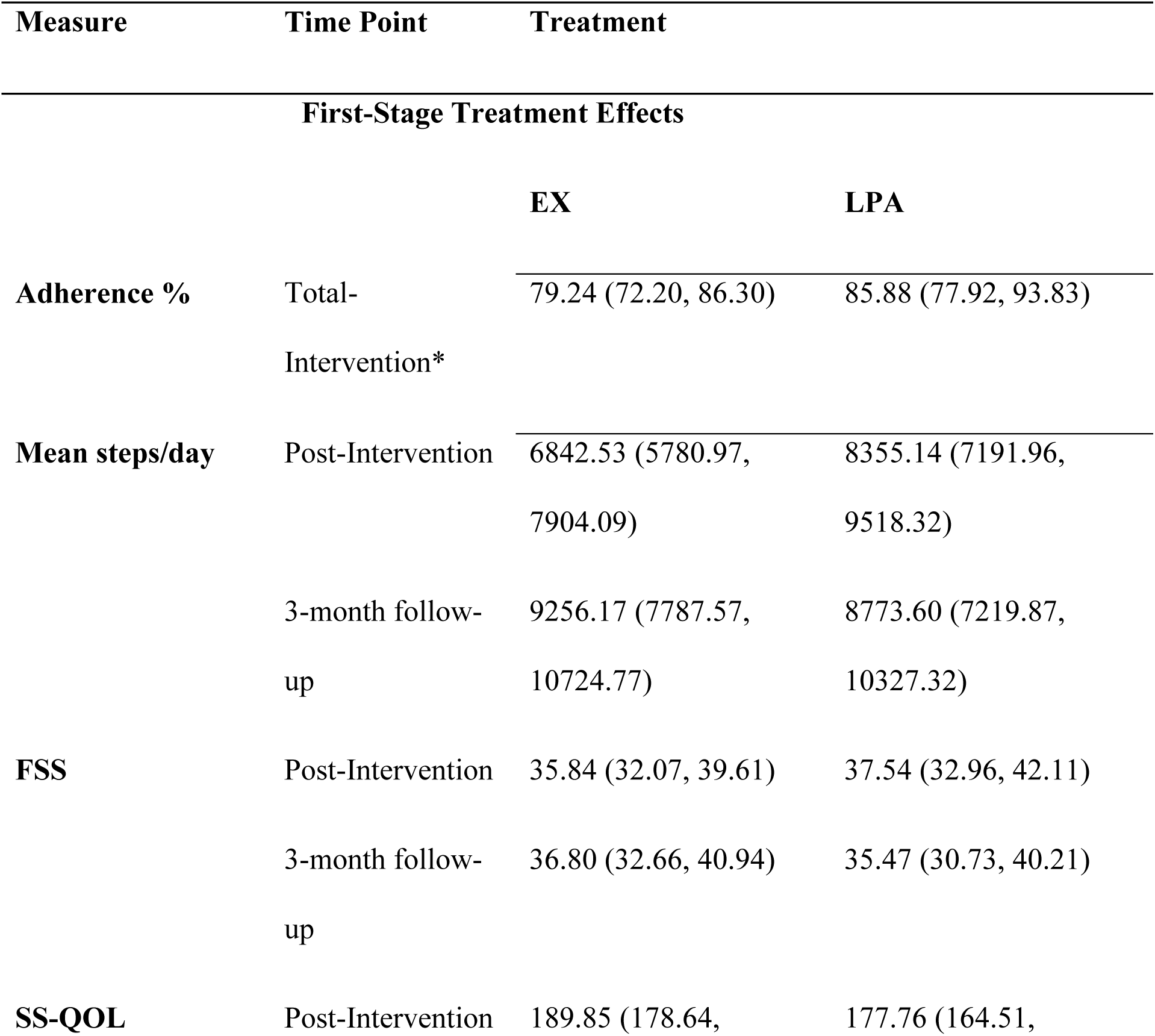
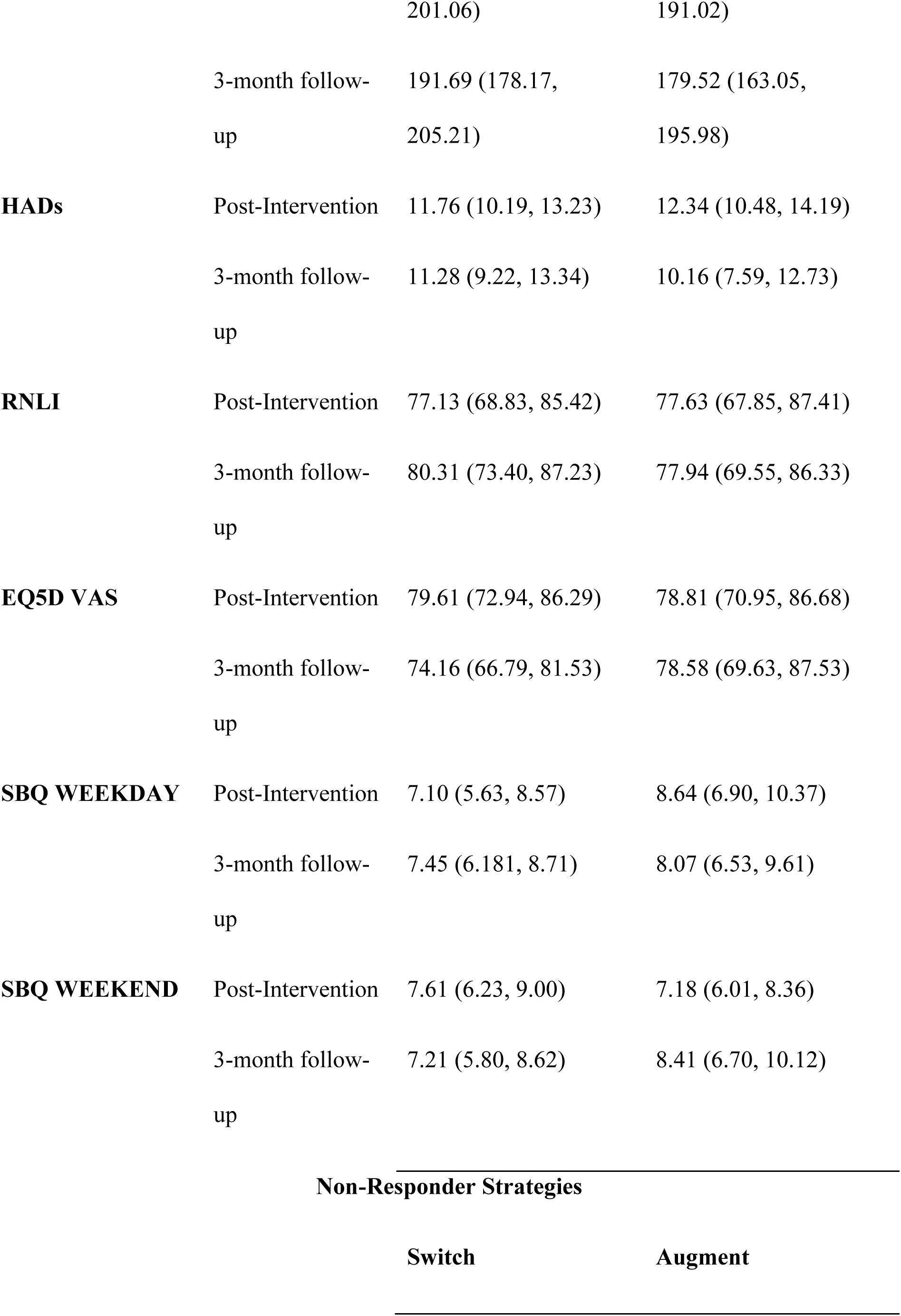
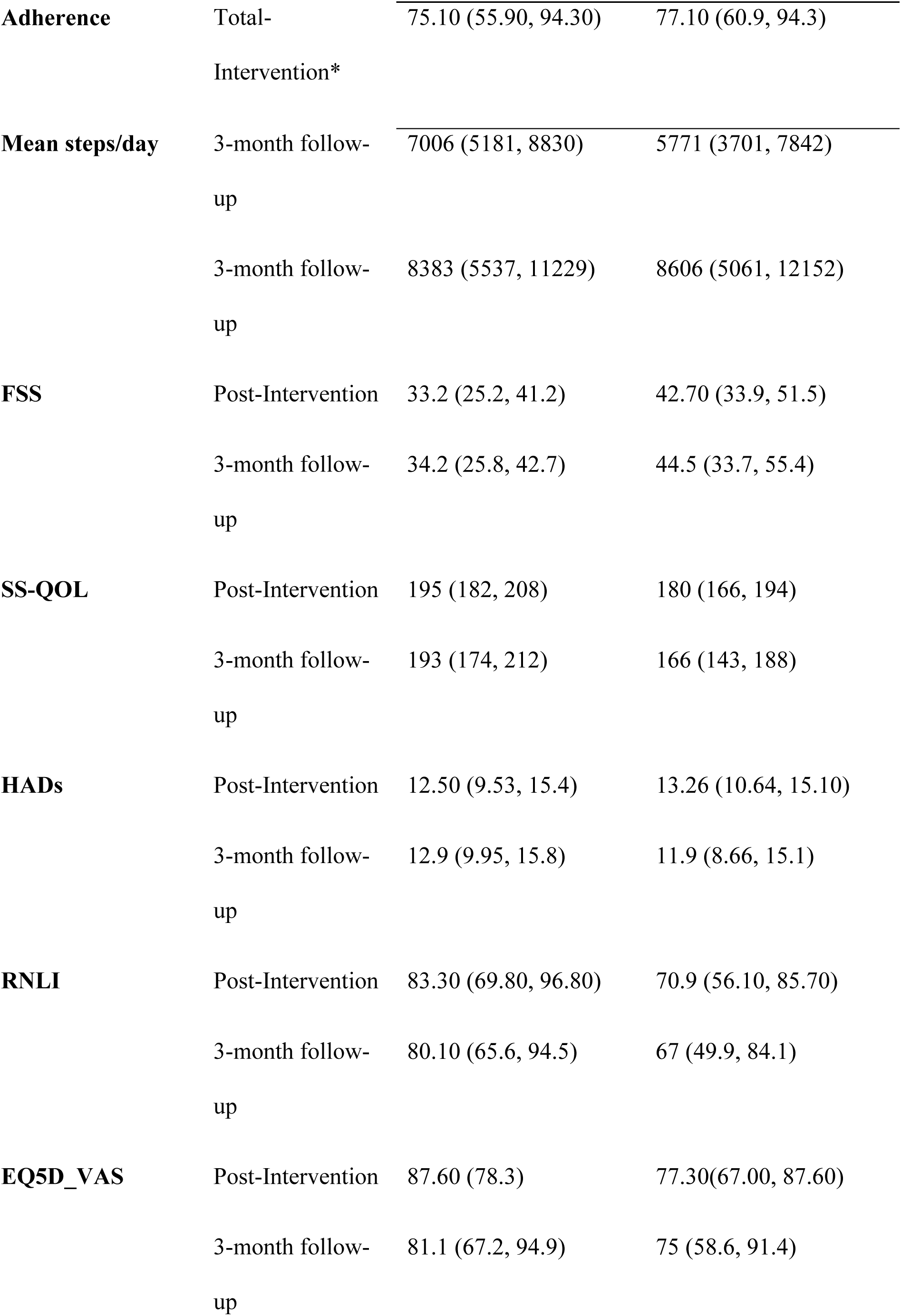
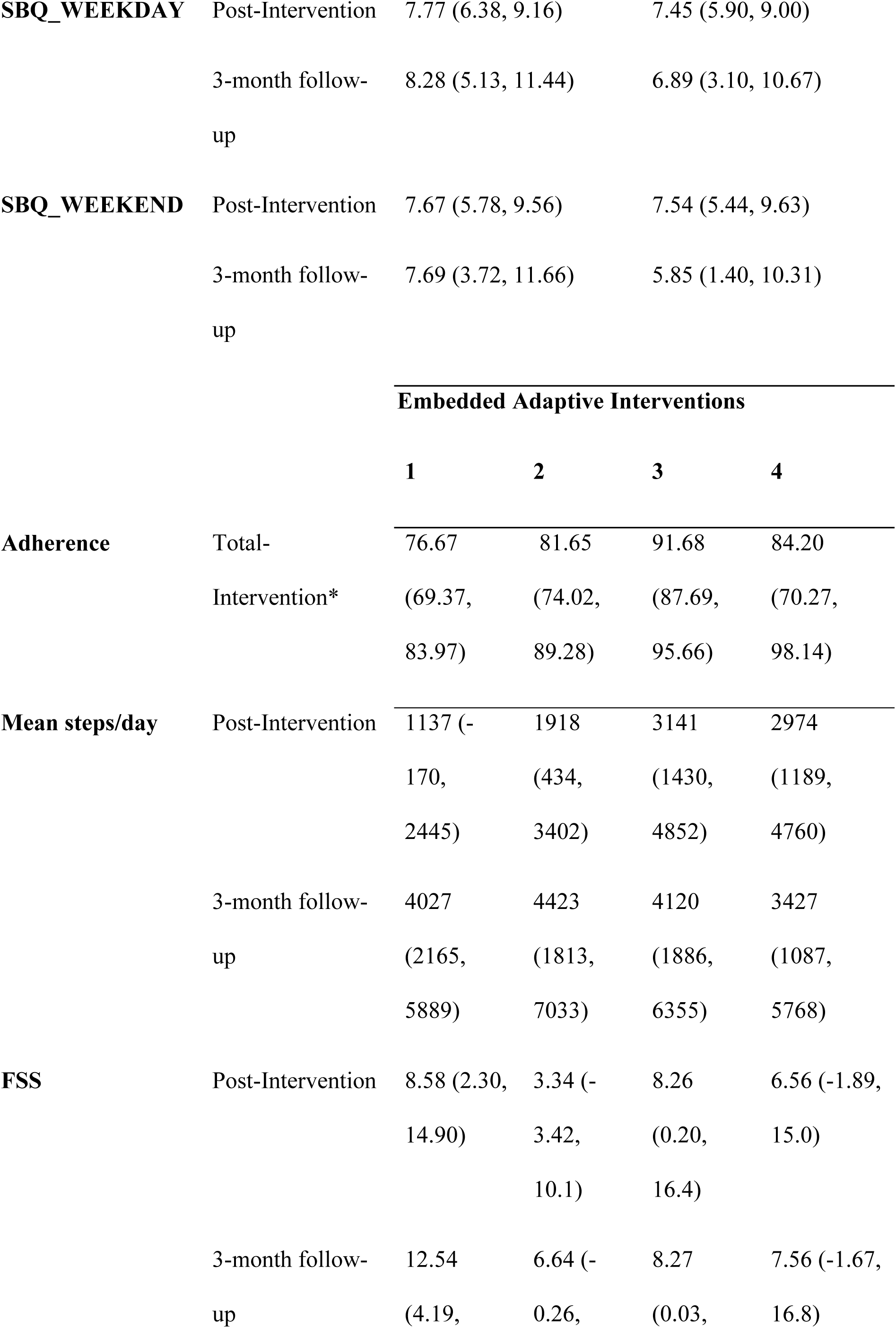
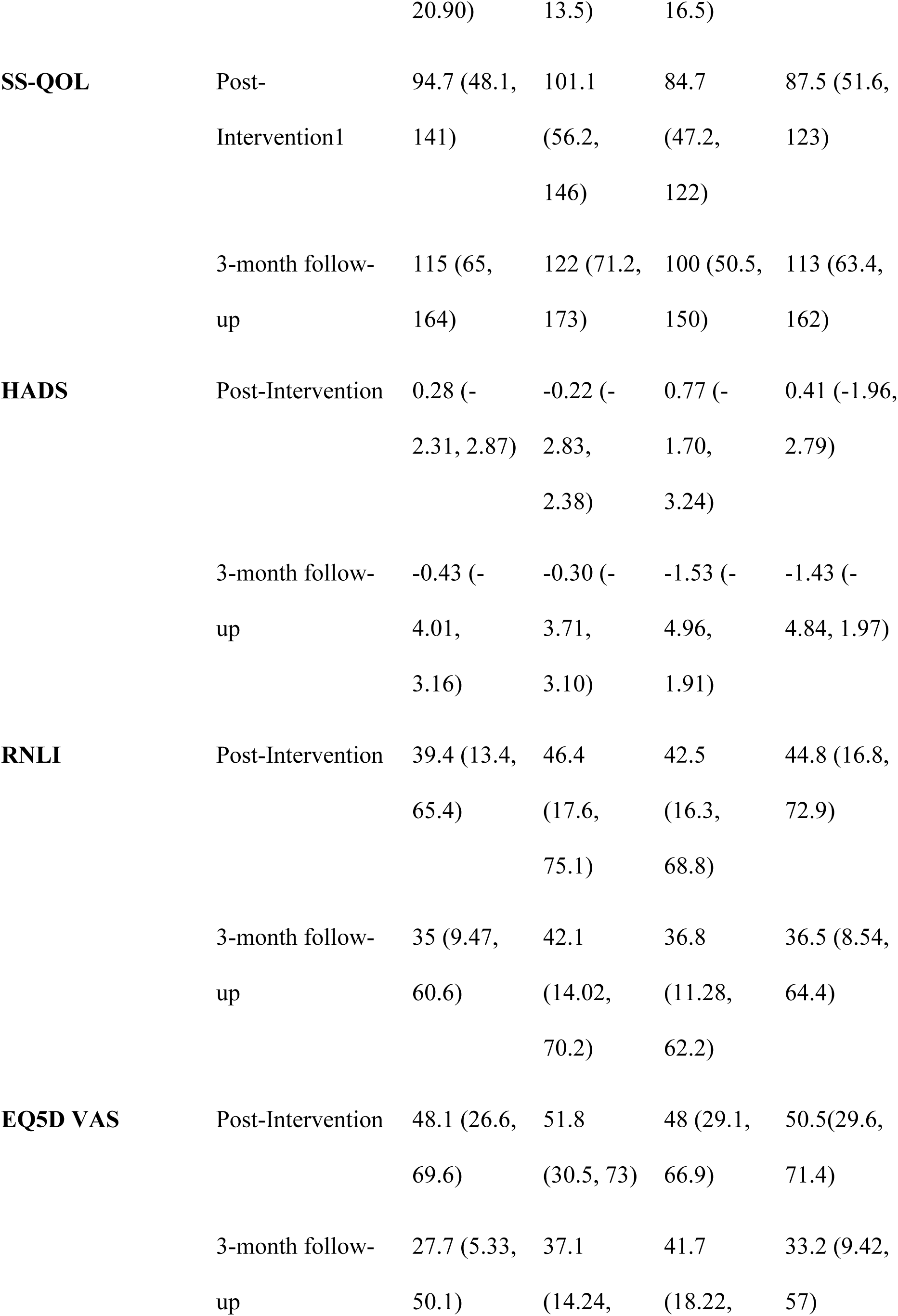
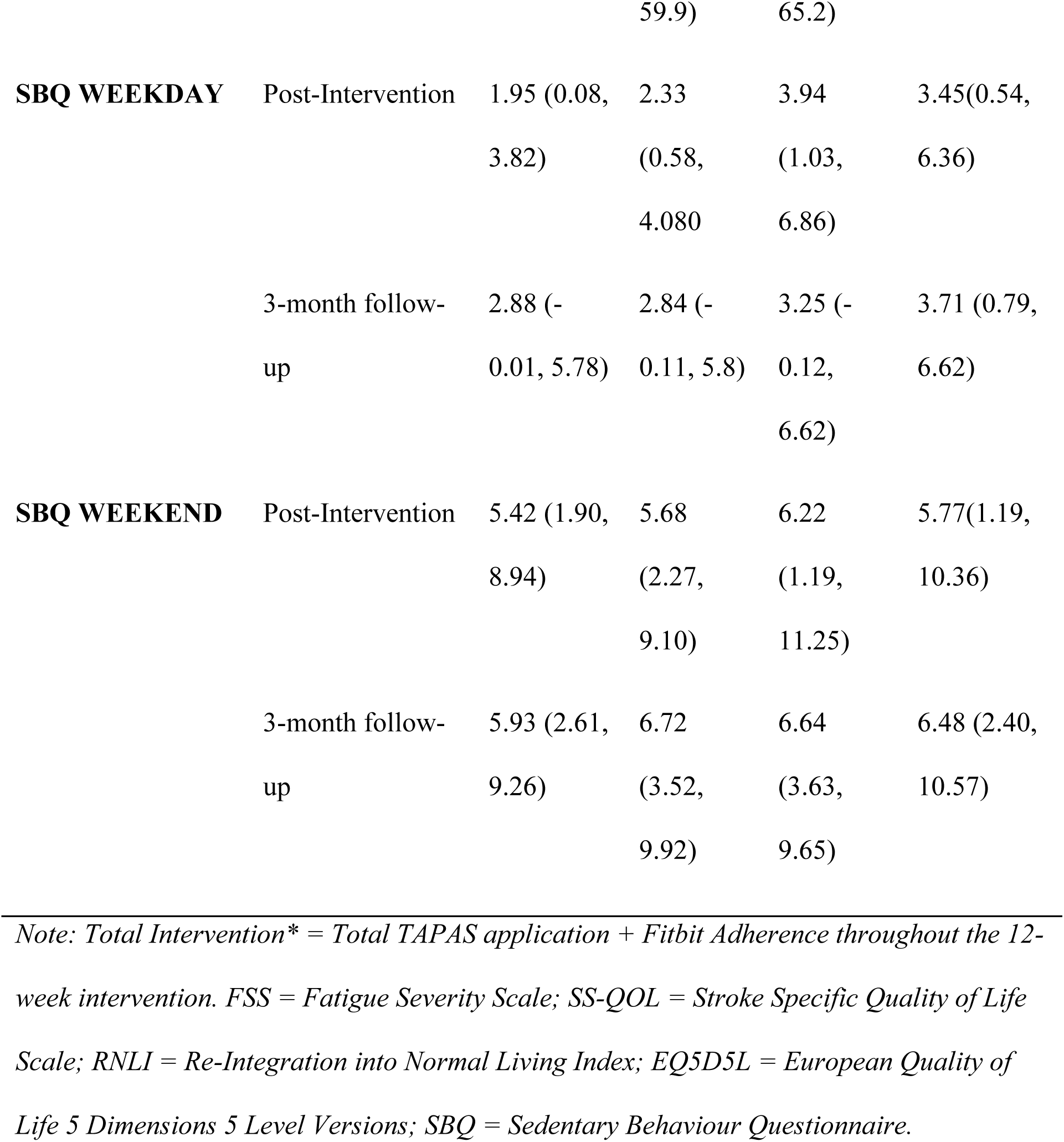
Marginal Means and 95% Confidence Intervals for SMART Secondary Outcomes.

### Secondary Outcomes

See Table 5 for the marginal means and confidence intervals of all secondary outcomes, controlling for baseline measures, related to comparing first-stage treatment assignment, non-responder strategies and EAIs. For initial treatment assignment (EX vs. LPA), no significant differences or trends were observed across secondary outcomes at post-treatment and 3-month follow-up.

Regarding non-responder strategies (switching versus augmenting treatment), no significant differences were observed across secondary outcomes, controlling for baseline outcomes and first-stage treatments. However, marginal means indicated positive trends favouring switch strategies in terms of mean steps/day, fatigue, stroke-specific quality of life, general quality of life, and re-integration into normal living at post-intervention. At the 3-month follow-up, positive trends favouring switch strategies were observed for fatigue, stroke-specific quality of life, general quality of life, and re-integration into normal living. Positive trends favouring augmenting treatment was observed for sedentary behaviour.

Evaluating the four EAIs at post-intervention using weighted and replicated regressions showed positive trends in mean steps/day for EAI3 – first-stage intervention LPA with a non-response strategy of augment with EX, and EAI4 – first-stage intervention LPA with a non-response strategy of switch to LPA (see Table 1). EAI2 - first-stage intervention EX with a non-response strategy of switch to LPA, showed positive trends in fatigue, stroke specific quality of life, psychological distress, and re-integration into normal living. EAI1-first-stage intervention EX with a non-response strategy to augment with LPA, and EAI2 - first-stage intervention EX with a non-response strategy of switch to LPA, showed positive trends in reducing sedentary behaviour. At the 3-month follow-up, positive trends were shown for EAI2 - first-stage intervention EX with a non-response strategy of switch to LPA, related to mean steps/day, fatigue, stroke-specific quality of life, and re-integration to normal living.

EAI3 - first-stage intervention LPA with a non-response strategy of augment with EX, showed a positive trend related to general quality of life.

## Discussion

The aim of this study was to examine the feasibility of an adaptive, mHealth PA intervention for community-dwelling people with stroke, using a SMART design. Trends in clinical efficacy by comparing first-stage intervention components, second-stage non-responder strategies and trends in primary and secondary outcomes across four EAIs, were also examined.

The TAPAS feasibility SMART has provided the methodological and statistical data needed for the development of a full-scale SMART in PA post-stroke. The feasibility indicators suggest that a definitive SMART is viable with minor design modifications that are discussed throughout, to enhance implementation and clinical integration. Eighty-five percent of the people who expressed interest in the study started the TAPAS intervention, which reflects a higher recruitment proportion compared with other digitally-delivered trials in stroke rehabilitation (40, 41). The TAPAS cohort included a relatively high proportion of male participants, with a relatively low mean age of 51 years, mild-to-moderate self-reported disability, relatively good health-related quality of life, familiarity with mobile application use and relatively high levels of PA at baseline. Systematic reviews and meta-analyses have consistently demonstrated low levels of PA among people post-stroke (42, 43). Fini et al (2018) (42) outlined that people in the subacute phase post-stroke showed a mean of 5,535 steps/day, whereas those in the chronic phase demonstrated mean steps/day of 4,078 (42). In comparison, the TAPAS participants demonstrated a baseline mean daily step count of 7,698.

This relatively high level of PA among participants is not unexpected, given the common recruitment bias in PA interventions toward people who are already physically active (44). There appears to be a degree of sociodemographic under-representation, particularly among female participants, people with lower education attainment, people who are older (only 18% of the TAPAS population was over 65 years) and those who are less independently mobile post-stroke. This may be attributable to well-documented challenges in recruiting diverse populations into digital interventions targeting chronic health conditions (45). The definitive SMART should consider more target recruitment strategies to enhance the external validity of findings.

High retention rates (87% at post-intervention and 80% at 3-month follow-up) were demonstrated in the TAPAS cohort, comparing favourably with the existing evidence base. A recent study examining the feasibility and acceptability of a community-based PA intervention among people with stroke (n=19) demonstrated 65% recruitment rate and 59% retention (46). However, the potential for mHealth to augment retention is an emergent theme in the literature post-stroke. Thurston et al (2025) (41) conducted a two-arm trial among 114 people with transient ischaemic attack or mild stroke, examining the feasibility and acceptability of a mHealth-delivered PA intervention. Results demonstrated very high retention rates, by using digital questionnaires (98%), sensor-derived PA (92%) and blood pressure monitoring (97%). Recruitment and retention outcomes were robust, indicating the effectiveness of the participant engagement strategies employed. Over a five month-period, a total of 50 participants were successfully enrolled in the study. These outcomes are likely attributable to the implementation of a multi-faceted recruitment approach encompassing direct engagement, individualised follow-up and patient-centred retention initiatives. In response to observed declines in attendance, targeted virtual follow-up interventions appeared to enhance adherence. The high retention rate achieved underscores the potential value of sustaining these strategies in the full-scale SMART.

Adherence to the TAPAS intervention was comparable (82%) with the findings of Thurston et al (2025) (41) wherein authors reported an adherence rate ranging from 76% to 96%, depending on the intervention component, for example, exercise sessions or counselling calls. The TAPAS trial is the first study in stroke to use the SMART design to empirically examine adherence rates among various EAIs throughout the trial. While adherence was very high across all EAIs (77%-92%), results demonstrated that EAI3 and EAI4 showed trends towards higher intervention adherence (92% and 84%, respectively). EAI3 and EAI4 involved initial treatment allocation of LPA and subsequently, depending on the participants’ individual responses to the intervention, continue in the LPA intervention (EAI3 or EAI4) or augment with EX (EAI3) or switch to EX only (EAI4). Higher adherence to EAIs that begin with first-stage LPA-underpinned by the COM-B model (28) of behaviour change, may highlight the value of integrating behaviour change strategies early in a future full-scale SMART to support treatment adherence. Nevertheless, the ability to maintain high adherence across different EAIs supports the feasibility of delivering structured virtual PA interventions among people post-stroke living in the community. The ongoing TAPAS process evaluation will also provide further detail on the acceptability of the interventions while highlighting areas for refinement.

This study represents the first SMART to examine the feasibility of adapting or modifying treatment strategies within a PA intervention post-stroke. Findings suggest that the re-randomisation procedure itself seemed to play an important role in the response of the participants who were deemed to be non-responders at six weeks. We noted trends toward an improvement from baseline in mean steps/day at post-intervention and 3-month follow-up among non-responders who switched interventions at six weeks, as opposed to augmenting their initial intervention. Similarly, positive trends were observed across other secondary outcomes among non-responders for switching treatments as opposed to augmenting their initial treatment allocation. For example, improvements were noted in stroke-specific quality of life and re-integration into normal living demonstrating the potential benefits of the EAIs that started with structured exercise (EX) and switched to LPA (EAI2). As noted in our systematic review of SMARTs used to develop PA interventions (17), few of the included studies also employed multiple strategies for non-responders at second stage, including augmenting and switching (47, 48, 49, 50, 51, 24). While switching appears to be a key non-response strategy for a future full-scale SMART, unexamined factors like participant preference may also warrant consideration as a non-response strategy.

Thompson et al (2024) (11) completed a randomised controlled trial among 250 people aged 21-85 years, walking without physical assistance post-stroke of ≥6 months duration, which were randomly assigned to a 12-week high-intensity walking intervention (FAST), a step activity monitoring behavioural intervention (SAM), or a combined intervention (FAST+SAM). Results demonstrated an improvement in mean steps/day found in the SAM (1542) and FAST+SAM (1307) groups, which represents an increase of >30% of the steps normally taken by people with chronic stroke (42, 43). The TAPAS trial demonstrated comparable improvements across various EAIs. These changes in steps/day are almost double the typical changes that have been found with previous walking interventions in people with chronic stroke (11). To further contextualize these findings, a recent meta-analysis reported that each additional increment of 1,000 steps per day is associated with a 15% reduction in all-cause mortality (52). Thus, the gains in daily step count activity found in the TAPAS cohort (in particular EAI2 and EAI3 at 3-month follow-up), may have important implications for the health of people living with stroke.

### Strengths and limitations

Innovations in trial design and analysis have occurred to address the problems of poor trial efficiency in traditional randomised controlled trials (54). In the current stroke research climate, there are acknowledged challenges in conducting clinical trials of adequate power in a timely manner (54). The use of the SMART design has potentially an indispensable role in advancing future clinical stroke research and care.

In our review of 20 SMARTs of PA interventions across a variety of populations (17), we noted that clearer rationale for the selection and timing of tailoring variables, and included measures is essential to advance PA SMART designs. A previous systematic review examining the quality of reporting of SMARTs (62) also noted methodological and reporting flaws among previous SMARTs. Of note, some features of SMART designs were seldomly reported, for example, inadequate reporting of the best EAIs (62). The TAPAS SMART has incorporated the recommendations of these systematic reviews, resulting in a methodologically rigorous approach to design, evaluation, and reporting. As shown in the current study, PA SMART designs should include at least two decision stages and use an objectively measured tailoring variable averaged over time to reflect the dynamic nature of PA. For a full-scale trial, the current design could potentially be improved by combining the objective PA data with participant-reported outcomes-e.g., participant satisfaction or acceptability. Including participant preference may also enhance real-word relevance, adherence, and treatment effectiveness of future full-scale PA SMARTs (17).

As described, EAIs are a set of sequential decision rules, each one corresponding to a key decision point in the patient’s history (53). An EAI represents a formalisation of the multi-stage and dynamic decision process potentially followed by healthcare professionals, which is the key concept of precision medicine. In this sense, identifying an optimal EAI would be a way to put evidence-based precision medicine into practice in stroke rehabilitation. To this end, the TAPAS SMART has identified important data on EAIs for PA promotion post-stroke which will inform the intervention components tested in the full-scale SMART.

While we did not have the statistical power to detect meaningful change in the primary or secondary outcomes; nonetheless, the feasibility findings presented, in addition to the magnitude of improvements in secondary outcomes, will inform the design of a full-scale SMART and targeted analyses of these important factors. Another limitation relates to the smartphone application which we designed to implement the intervention. Adaptations in technology intervention are dynamic and must be implemented quickly and an application with artificial intelligence could automate and individualize the adaptive process to increase PA (63). A process evaluation is currently underway to further examine the acceptability and feasibility of the TAPAS design and intervention among stakeholders.

### Conclusions

Although the benefits of a healthy lifestyle, including adequate PA and vascular risk factor control are well documented (64, 65), risk factors remain poorly controlled among people post-stroke (66). Recurrent stroke poses a significant threat to public health. The findings of this SMART provide first-in-class empirical information on the feasibility and trends in clinical efficacy of an adaptive PA intervention delivered via mHealth for people post-stroke. Results suggest that a definitive SMART of this intervention would be feasible. Findings contribute to the evidence on how best to develop and deliver mHealth PA interventions post-stroke, maximising efficacy.

## Data Availability

The data that support the findings of this study are available from the corresponding author upon reasonable request.

## Acknowledgments

We would like to acknowledge the input from all of the participants in the TAPAS trial.

## Non-standard Abbreviations and Acronyms

PA: Physical activity
FAST: High intensity walking program
SAM: Step activity monitoring behavioural intervention
EAI: Embedded adaptive intervention
SMART: Sequential, Multiple Assignment, Randomised Trial
EX: Structured exercise
LPA: Lifestyle PA
BCW: Behaviour Change Wheel
SBQ: Sedentary Behaviour Questionnaire
SS-QOL: Stroke Specific Quality of Life scale
HADS: Hospital Anxiety and Depression Scale
RNLI: Re-Integration into Normal Living Index
CASP: Cognitive Assessment Scale for Stroke Patients
GEE: Generalized estimating equation
PPI: Public and patient involvement

